# Translating a novel wildfire smoke exposure chamber system from lab-based experiments to community-engaged activities

**DOI:** 10.64898/2026.03.06.26346761

**Authors:** Lilian Liu, Shirley Ching-Hsuan Huang, Alison Hirata, Ione Jones, Ningrui Liu, Jeffry Shirai, Christopher Zuidema, Elena Austin, Edmund Seto

**Affiliations:** Department of Environmental and Occupational Health Sciences, University of Washington, Seattle, WA 98105, United States; Center for Environmental Health Equity, Department of Environmental and Occupational Health Sciences, University of Washington, Seattle, WA 98105, United States; Clean Air Initiative 2024, KHIMSTONIK, Toppenish, WA 98948, Yakama Nation Sovereign Lands; Division of Public Health, Larner College of Medicine, University of Vermont, Burlington, VT 05405, United States

**Keywords:** Wildfire smoke, PM_2.5_, low-cost air quality sensors, box fan filter, clean air delivery rate, indoor air quality, tent, community-engaged events

## Abstract

Wildfire smoke (WFS) events are an important public health concern for communities in the Pacific Northwest of the United States. Previous studies of portable air cleaners, including high efficiency particulate air (HEPA) filtration and do-it-yourself (DIY) box fan filters built with MERV 13-rated filters, have indicated that their use in residential settings may be an effective way to reduce indoor exposures to fine particulate matter during WFS episodes. The lower-cost, easy to build instructions and availability of materials of DIY box fan filters have made their distribution by both public health agencies and community groups an attractive approach to improve community preparedness. Here, we describe a low-cost, easy-to-assemble, portable exposure chamber system that can be used to support a variety of community-engaged demonstrations of WFS removal efficiency as well as provide a mechanism to estimate the efficiency of filtration systems in a controlled environment. We conducted experiments using the portable chamber to assess the clean air delivery rate (CADR) of a MERV 13-rated DIY box fan filter, which was found to be 92.2 and 145.2 cfm at low and high fan speeds, respectively. In addition to using the chamber system to evaluate the CADR of DIY box fan filters, we also provide a case-study example, working with a tribal community in Central Washington, who used the tent system for a live demonstration of a DIY box fan filter experiment during their community gathering to promote WFS and air quality intervention knowledge and distribution of box fan filters.

## 1. Background

Wildfire smoke (WFS) events are increasingly recognized as a concern for the Pacific Northwest (1), and events over the past decade have highlighted how poor air quality due to WFS can negatively affect human health (2,3). Washington state agencies, such as Public Health Seattle & King County (PHSKC) (4) and Washington State Labor and Industries (WALNI) (5), issue guidance and policies to protect residents and working populations from harmful health impacts. Adverse health outcomes are shown to be associated with acute WFS exposure, including exacerbation of symptoms in individuals with underlying conditions such as congestive heart failure (CHF), asthma, chronic obstructive pulmonary disease (COPD), and acute bronchitis (6–10). Cause-specific mortality and development of cardiorespiratory morbidities are also linked to chronic exposure to long-term WFS (3,11–14). Growing evidence also suggests negative mental health/well-being impacts (15–18), adverse pregnancy outcomes (19–23), metabolic disorders like Type II diabetes (10,24), and cognitive impacts (25–27) are linked to WFS exposure. Studies also suggest inequity in the impacts of WFS related to socioeconomic and other population vulnerability indicators (28–30), including increased risks for children and elderly populations, and disadvantaged, rural, and indigenous tribal communities (31–35).

To mitigate the adverse health impacts associated with WFS exposure, portable air cleaners (PACs) have been shown to be effective in improving indoor air quality by removing harmful WFS fine particulate matter, such as particulate matter that is 2.5 micrometers in diameter or smaller (PM_2.5_) when used properly (36–39). However, due to the cost of commercial PACs based on high-efficiency particulate air (HEPA) filtration, another commonly used indoor air treatment, do-it-yourself (DIY) box fan filters that utilize MERV 13-rated filtration, has become a popular alternative due to its relative low-cost and accessibility. A study conducted by researchers at the US Environmental Protection Agency (US EPA) found that in well-controlled experiments, DIY box fan filters can be as effective as branded PACs in PM_2.5_ particle removal (36). Additionally, DIY box fan filters can be constructed with various number of filters (i.e., 1, 2, or 4 (also known as Corsi-Rosenthal box air filters (40))) for different filtration needs. The lower price and customizable feature of DIY box fan filters could serve as a more economical and flexible option for low-income communities.

To compare particle removal efficacy of different PACs, the clean air delivery rate (CADR) metric is used to quantify the amount of clean air being filtered and delivered in a standard unit of time. The CADR can be determined experimentally, typically in well-controlled laboratory conditions such as the Association of Home Appliance Manufacture (AHAM) certified testing chamber. The AHAM exposure chamber, which has been employed by many PAC evaluation studies, is designed to simulate a small room with size of 29.3 m^3^ (41,42). This exposure chamber allows for standard testing conditions and reproducible air treatment efficacy testing studies (43). However, AHAM exposure chambers are not appropriate for all situations, especially due to their high cost, complexity, and lack of portability.

Ideally, a lower-cost, simpler to operate, and portable alternative to the AHAM exposure chamber would provide comparable WFS experimental results for PAC evaluation. This simpler and portable exposure system would greatly facilitate a variety of use cases, including in scientific research, education and training, product development and testing, as well as for indoor air quality demonstrations at community-based events. For example, a simpler chamber design utilizing low-cost real-time continuous particle reading instruments might be more appropriate for use at community events, where the goal is to improve community knowledge and risk communication during WFS (44). The chamber could be used to demonstrate particle concentration decays as a PAC operates to help members of the community understand how to use a PAC and its efficacy in removing particles from indoor air – to *show*, rather than *just tell* community members the utility of portable air filtration. Indeed, others have described the importance of increased community-based education and community response capacity as ways to reduce residents’ smoke exposure during WFS events from a public health perspective (44). While factsheets, pamphlets, and various social media content such as video clips are useful in disseminating public health guidance to improve people’s awareness and knowledge about WFS and its impacts (45–47), live demonstration of research in community-based settings has been shown to be a favorable strategy to promote health behavior change (48–50). Therefore, transferring the AHAM lab-based particle removal evaluation of DIY box fan filter to a simpler approach that can be used in a community-based setting would be a good option to bring awareness and promote knowledge of WFS exposure, its health impacts, and intervention strategies.

The objective of this paper is to describe a low-cost, easy-to-assemble, portable exposure chamber system for WFS experiments in both laboratory settings and community-engaged demonstrations. Our specific application of the exposure chamber system was to visualize and evaluate the effectiveness of DIY box fan filter particle removal efficacy. We first utilized the system to assess the clean air delivery rates (CADRs) of the DIY box fan filter. We then worked with a tribal community organization to use the system for a live demonstration to promote clean air knowledge and behavioral change for future WFS episodes.

## 2. Methods

### 2.1 Tent exposure chamber and woodsmoke experiment setup

Our low-cost portable exposure chamber consists of a large instrumented portable tent. We used a commercially available horticultural grow tent for its modular assembly and durability. A 333 ft^3^ (9.43 m^3^) canvas grow tent (VIVOSUN S105 10x5 Grow Tent) was purchased through online retailer Amazon at $289.99 USD. Briefly, the tent exposure chamber is built of Oxford canvas with a mylar inner layer and metal framework, with duct tape at openings for leakage prevention. Detailed information on the tent exposure chamber can be found in Supplementary Information Section 1.

The tent can serve as an exposure environment for particles generated from various sources. For our experiments focused on WFS, for particle generation, we utilize a food-grade smoke gun (NEZActive Cocktail & Food Smoker smoking gun; ASIN: B096M954P1) and woodchips (Breville Classic Smokehouse Wood Chip; ASIN: B07168H4SG). Using the smoke gun to burn 1 teaspoon of woodchips, a bolus of woodsmoke is introduced in the tent exposure chamber, using a resealable inlet at the zipper port at the back of the tent. The inlet is closed once woodsmoke introduction is complete (Fig. 1). The woodsmoke generation protocol is described in detail in Supplementary Information Section 2. A pedestal fan (Lasko Adjustable-Height 16 in. 3 Speed White Oscillating Pedestal Fan) inside the tent is used to promote mixing and uniform particle concentration.

**Figure 1.**
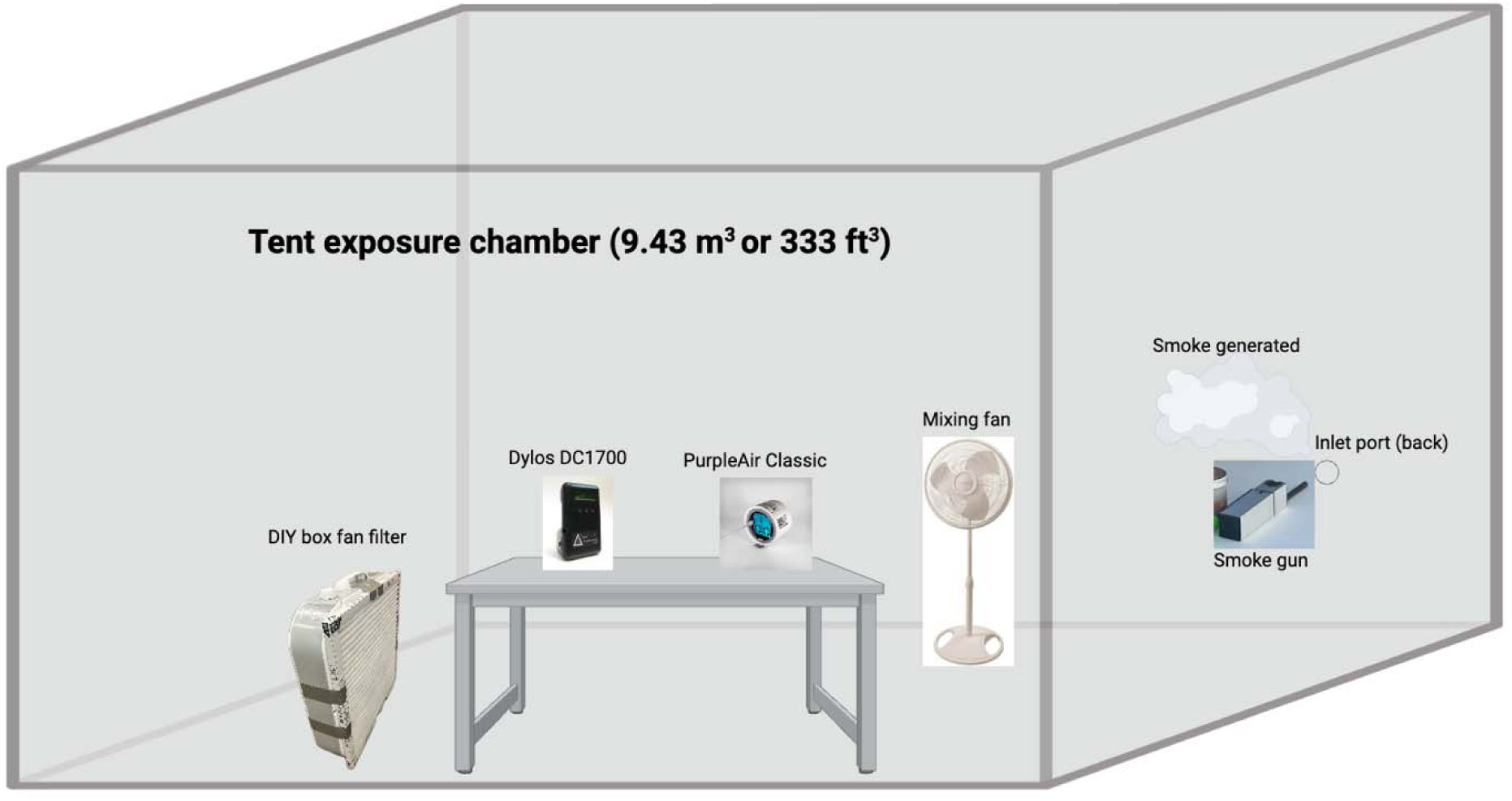
Schematic diagram of the exposure chamber and experiment set up. Smoke gun introduces woodsmoke from the outside of the chamber through the inlet port. Particle sensor TSI 3330 (not displayed) is outside of the tent, but sampling air from within the tent.

During the lab-based CADR experiment, we used three different particle measurement instruments to assess the particle concentration for data analysis and results comparison. First, a real-time continuous research-grade optical particle sizer (TSI optical particle sizer 3330; TSI Inc.), which was stationed outside of the tent exposure chamber with the instrument’s air inlet sampling particle concentration within the tent using a static-free sampling tube through an inlet of the tent. The TSI 3330 was set to the predefined protocol ASHRAE 52.2 (51), with 13 size channels between 0.3 to 10 μm in diameter on TSI 3330 with the particle density of 1.98 g/cm^3^. Second, a low-cost particle instrument (PurpleAir Classic; PurpleAir) was used to monitor the particle concentration at 1-second intervals in the tent exposure chamber. Because the instrument does not rely on a conventional sampling pump and tubing, the instrument was placed within the tent to directly sample the air. The PurpleAir Classic, which contains two separate particle sensors, was not calibrated. We modified the instrument, following the manufacturer’s guidance (52) to customize the sensor data logging rate from 2-minute to 1-second to match the data logging rate of TSI 3330. As described below, the TSI 3330 measurements were used to calibrate the Purple Air measurements for the particle source used in this experiment.

Third, we utilized another low-cost particle instrument (Dylos DC1700 Air Quality Monitor; Dylos corp.). The Dylos also does not utilize a sample pump or tubing, and thus, the instrument was placed within the tent. The Dylos collects particle concentrations every 10-seconds. The Dylos serial port was connected via a serial-to-USB cable to a PC laptop situated outside of the tent, which was used for data logging and to view particle concentration in real-time during the experiment.

### 2.2 Lab-based DIY box fan filter characterization using low-cost particle sensor

We followed standardized instructions for building a DIY box fan filter (53). In short, a MERV 13-rated filter was secured to the back (input side) of a Lasko 20-inch 3-speed box fan (Fig. 2) with duct tape. To test the CADR of the DIY box fan filter, we performed the box fan face velocity measurement with MERV 13 filter attached under ambient indoor environment followed by the in-tent particle removal experiment.

**Figure 2.**
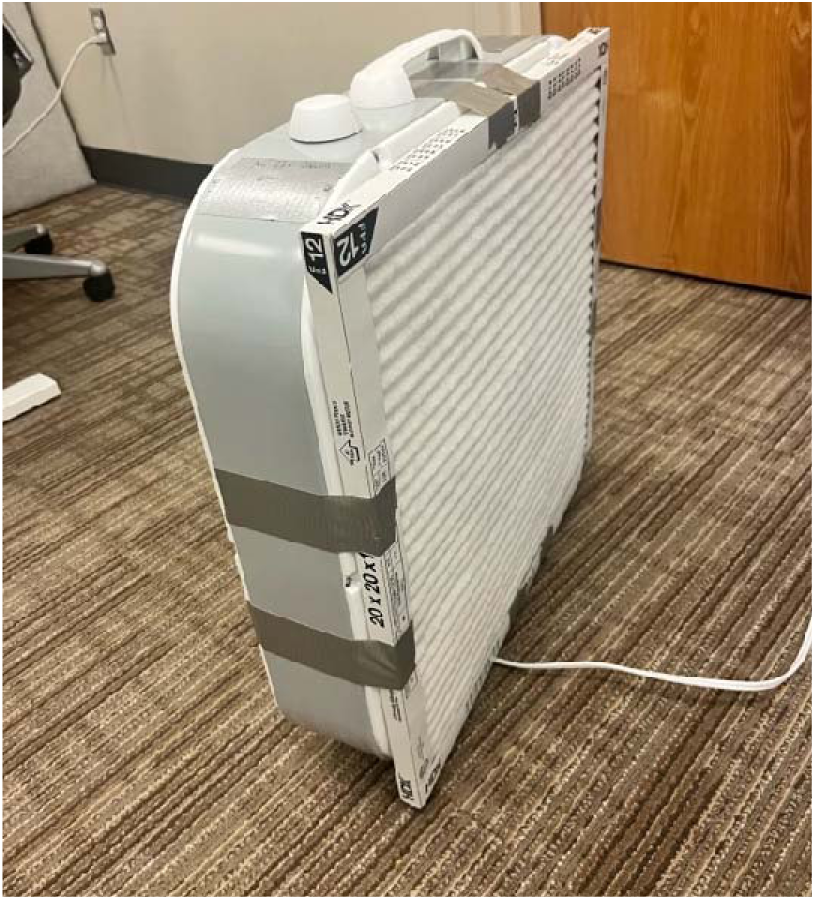
Photo of the DIY box fan filter under the ambient indoor environment condition.

To assess air flow rate through the filter (Eq. 1), we measured the face velocity by collecting repeated hot-wire anemometer (TSI VelociCalc Multi-Function Ventilation Meter 9565; TSI Inc.) readings following a grid pattern (i.e., dividing the box fan surface equally into nine square grids) along the face of the fan 6 inches away from the inlet side. We repeated the measurement three times for each grid at each fan speed (i.e., low, medium, and high). Air flow rates for each fan speed were calculated as the average value from individual repeated measurements among 27 measurements (a total of 81 measurements across all fan speeds). Detailed fan face velocity measurements can be found in Supplementary Material section 3. DIY box fan filter face velocity testing.

Box fan air flow rate (cfm) was calculated using Equation 1:

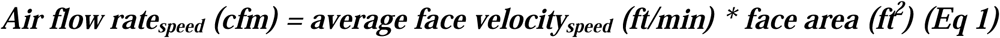

where the average face velocity was the average of the measurements at a specified box fan speed (i.e., low, medium, or high). The face area of the fan was obtained from the product website (16 x 20 inches).

To calculate DIY box fan filter’s CADR, which is a measure of air purifier’s air pollutant removal effectiveness, we introduced woodsmoke to the tent exposure chamber and measured the decay rates using TSI 3330, PurpleAir Classic, and Dylos air monitors when the DIY box fan filter was set to different fan settings. We used the TSI 3330 as our reference instrument for its lab-grade accuracy and decided on an initial target particle concentration within the tent of 100 μg/m, which represents an “unhealthy” level based on the air quality index (AQI) (54). We followed the woodsmoke generation protocol described in 2.1 and performed one round of “natural decay” with the box filter fan off to account for deposition and leakage of the tent exposure chamber and three repeated measures for each box filter fan setting. For both the natural decay experiment and the experiments for different box fan filter speeds, once the particle concentration target was achieved after introduction of woodsmoke, we zipped the tent shut and waited for the concentration to decay back to a background level of around 10 μg/m^3^. We then turned off the DIY box fan filter, completing an individual experimental trial. We performed one trial to measure the natural decay constant and three trials for each fan speed. Exponential decay constants were assessed for each trial (Equation 2):

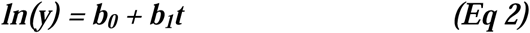

where y is the particle concentration, t is the elapsed time in hours, b_0_ is the natural logarithm of the initial peak background concentration, and b_1_ indicates the decay constant of a trial. We obtained the decay constant for the natural decay b_nat_ and experimental decays b_exp_ for each experimental trial using the decay calculation. The net decay coefficient was computed as the difference between the experimental decay for a particular fan speed and the natural decay (Equation 3):

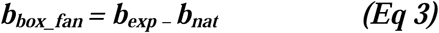

We also followed previous work from Huang et al., to calculate CADR at low, medium, and high speed, respectively (55) (Equation. 4):

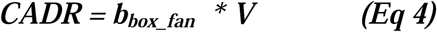

where CADR in units of m^3^/h is calculated for DIY box fan filter particle removal efficacy testing. In the equation, b_box_fan_ is the decay coefficient (per hour), which was calculated in Equation 3, and V is the total volume (m^3^) of the tent exposure chamber. All analyses were conducted in R (version 4.3.1), using the packages data.table (56), tidyr (57), tidyverse (58), finalfit (59), flextable (60), officer (61), ggplot2 (62), ggpubr (63), and viridis (64).

### 2.3 WFS community-engaged event

After conducting laboratory evaluations of the DIY box fan filter, the exposure tent system was used to demonstrate the use of box fan filters at a community event where the box fan filters were being distributed. As context, the University of Washington (UW), Center for Environmental Health Equity (CEHE) was funded by US EPA Thriving Communities Technical Assistance Center (TCTAC) program. The center aimed to provide technical assistance to tribal and community-based organizations in EPA Region 10 states (Alaska, Idaho, Oregon, and Washington) to strengthen their capacity and effectiveness in applying to and successfully managing environmental and energy justice grants and programs. In fall 2024, a Tribal nonprofit organization in Central Washington reached out to CEHE to request technical assistance on WFS resources with exposure mitigation strategies (e.g., information handouts) at a community-engagement event they were hosting. Upon exchanging ideas, the group expressed enthusiasm in having a live demonstration using the tent exposure system to showcase the usage of DIY box fan filters to reduce indoor WFS exposure to support their DIY box fan filter giveaway effort. The organization serves a community that frequently experiences WFS impact (65–67). To support their Clean Air community event where the community leader planned to distribute DIY box fan filter, we prepared a short video demonstrating DIY box fan filter air cleaning efficacy in the tent exposure system prior to the community-based event. Additionally, to aid in wildfire smoke and air quality information sharing, we also prepared handouts on Wildfire Smoke and Air Quality in the region by gathering existing information from peer-reviewed publications (39,65,68) and evidence-based online resources such as US EPA (69–71), UW Department of Environmental and Occupational Health Sciences (DEOHS) (53), and Centers for Disease Control and Prevention (CDC) (72). Both the video and handouts are now publicly available on YouTube (73) and UW CEHE website (74). The handouts are also provided in Supplementary Information section 4.

Approximately 100 people attended the event. Many attendees have strong ties to the community, and shared that they lived in the area for many years, if not their whole lives. Most attendees were middle-aged and expressed deep concerns about environmental health and WFS episodes. The event hosted performances and presentations from local art groups and state agencies to share their cultural heritage and history and promote air quality education.

### 2.4 DIY-box fan filter particle removal live demonstration

The community event organizer and CEHE outreach manager, along with other community members, planned the logistics for using the exposure tent for a live demonstration, which included identifying a suitable outdoor location for the demonstration. It was determined that the live demonstration should use the tent exposure system to show community members the effectiveness of the DIY box fan filter in removing smoke from indoor spaces. During the allotted time on the day of the event, we set up the tent outside of the event venue (Fig. 3) and conducted a DIY box fan filter trial similar to the protocol described in 2.2. To keep the demonstration manageable, we only used the Dylos DC1700 particle counter to monitor the woodsmoke particle concentration inside the tent. The benefit of using the Dylos DC1700 (as opposed to the PurpleAir) is that it allows users to observe the current particle count concentration on its LCD screen. However, the Dylos LCD display does not display a time-series of the changing concentration as a tent experiment is conducted. A time-series visualization can be helpful for community events, and demonstrations of the effect of air cleaners, as the time-series would show particle decay with time. This was solved by utilizing the serial port interface of the Dylos that allows it to be connected to a laptop computer for off-device data logging and visualization. By connecting the Dylos to a laptop, we were able to run R scripts to facilitate real-time visualization of the particle concentration as a time-series for the decay experiments. The first R script is responsible for data-logging the particle concentrations to a comma-separated-value (CSV) file on a connected laptop. The second R script reads the saved CSV file to continually refresh a plot of the concentration time-series. Both scripts are provided in Supplemental Information.

**Figure 3.**
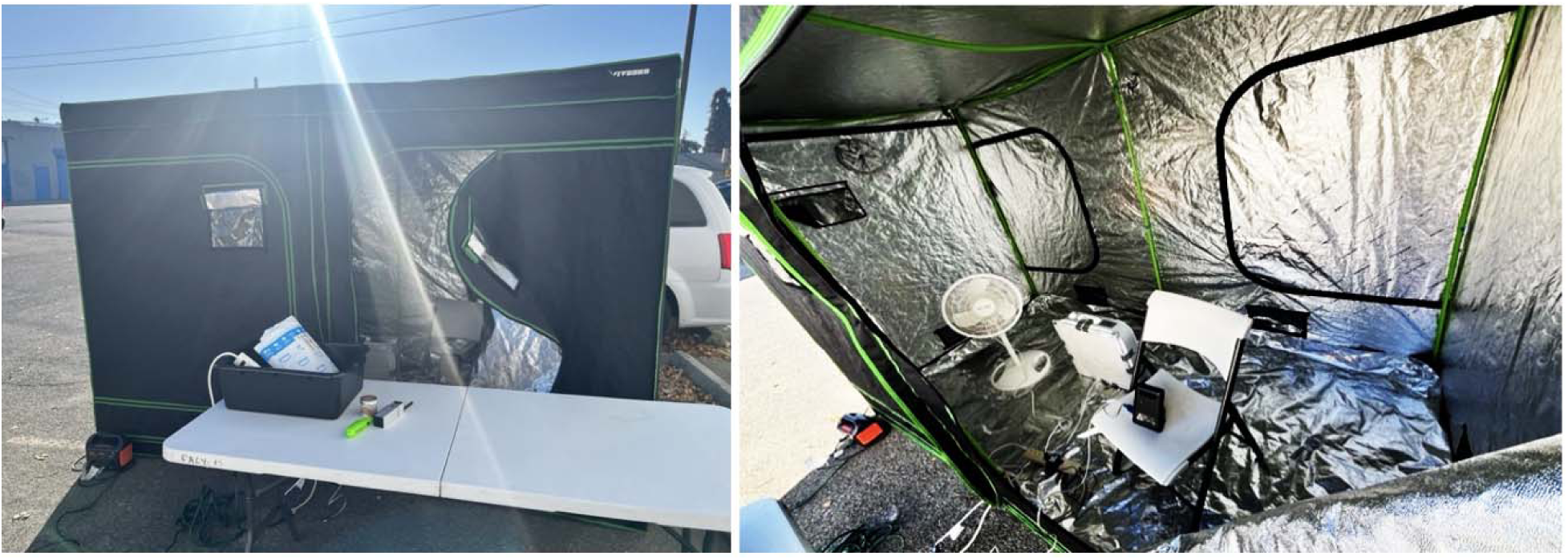
The tent exposure chamber and experiment material at the community event in Washington (left). Inside the tent exposure chamber (right) during the particle removal experiment at the community event in Washington where Dylos was placed on a chair inside the tent exposure chamber to monitor particle concentration in real-time; next to the chair on the tent floor was the DIY box fan filter and the standing mixing fan which were used to remove particles and promote air mixing, respectively. A laptop (not shown) wa connected via serial cable to the Dylos and placed on the table outside of the tent to show changing particle concentration as a real-time updating time-series plot.

During the live demonstration, we gave a short introduction to the experiment and invited community members to engage by assisting in introducing the woodsmoke into the tent. Using the Dylos particle concentration readings and connected laptop time-series visualization, attendees observed the particle concentration increase after introducing woodsmoke into the tent exposure chamber and decrease when the DIY box fan filter was turned on at medium speed. Attendees were encouraged to share their thoughts and questions throughout the live demonstration. No community members were allowed to go inside the tent at any point during the demonstration.

## 3. Results

### 3.1 CADR results of the DIY box fan filter

#### 3.1.1 DIY box fan filter fan characteristics

In the analysis, we focused on results from TSI 3330 due to its accuracy, and the PurpleAir Classic due to its wide use in real-world settings. Results from Dylos can be found in Supplementary Information sections 5 and 6. The DIY box fan filter had the face velocities of 55.5 ft/min, 86.3 ft/min, and 101 ft/min at low, medium, and high fan speeds, respectively. The air flow rates at low, medium, and high fan speeds were 741 cfm, 1150 cfm, and 1350 cfm (Table 1).

**Table 1.** DIY box fan filter fan characteristics. SD and SE stand for “standard deviation” and “stand error” that accounted for uncertainties in repeated measures for each fan speed.

| Fan Speed | Face velocity | Face velocity | Face velocity | Air flow rate,<br>cfm |
| --- | --- | --- | --- | --- |
|  | Mean, ft/min | SD, ft/min | SE, ft/min |  |
| Low | 55.5 | 12.2 | 3.87 | 741 |
| Medium | 86.3 | 13.7 | 4.35 | 1,150 |
| High | 101.0 | 24.7 | 7.82 | 1,350 |

#### 3.1.2 Particle size distribution and concentration statistics

During the CADR experiment, we observed a particle size distribution with mostly smaller particles that included sizes in the 0.3 μm diameter size bin before the smoke introduction and after the particles were removed, (Fig 4, panel A and C), but more slightly larger particles in diameters of 0.4, 0.55, and 0.7 μm when woodsmoke was introduced into the tent exposure chamber (Fig. 4, panel B). This indicated burning woodchip released a larger number of bigger particles, possibly due to incomplete combustion (75,76).

**Figure 4.**
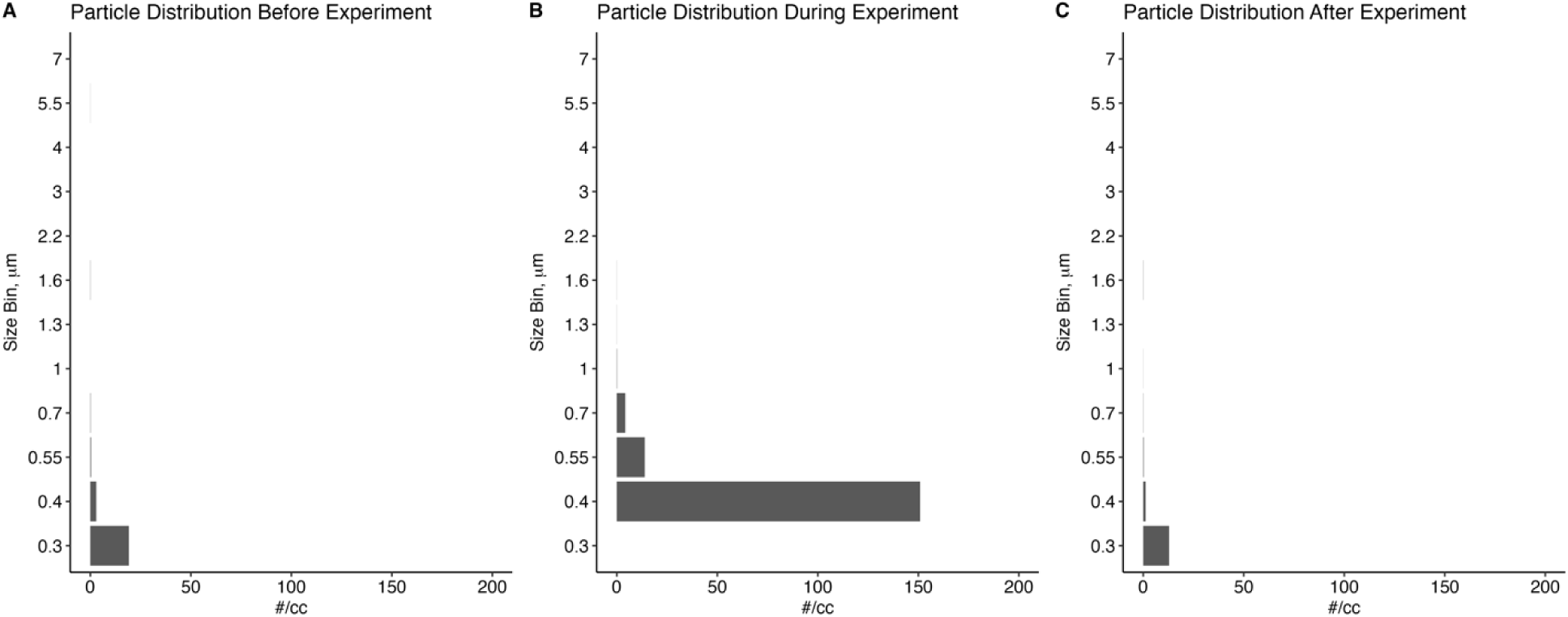
Particle size distribution before, during, and after one CADR experiment trial.

Particle concentrations during decay experiments ranged from 0.8 μg/m^3^ to 91.9 μg/m^3^ with a mean concentration of 31.5 (SD= 18.7) μg/m^3^ (Table 2). Compared to TSI 3330 particle concentration distribution, particle concentrations ranged from 1.0 μg/m^3^ to 510.0 μg/m^3^ from PurpleAir Classic (Table 2), with a mean concentration of 175.2 μg/m^3^ (SD= 114.9 μg/m^3^). Overall, results from PurpleAir Classic were higher compared to TSI 3330 across all experimental trials.

**Table 2.** PM_2.5_ concentration (μg/m^3^) summary statistics during CADR decay experiment.

| Particle Monitor | Mean | SD | Median | Min | Max |
| --- | --- | --- | --- | --- | --- |
| TSI 3330 | 31.5 | 18.7 | 32 | 0.8 | 91.9 |
| PurpleAir Classic | 175.2 | 114.9 | 148 | 1.0 | 510.0 |

#### 3.1.3 Particle decay

To visualize the decay curves and compare decay results between TSI 3330 and PurpleAir Classic, we normalized the particle concentration by using particle concentration divided by the max particle concentration for each decay experimental trial, respectively. After normalizing the particle concentration (Fig. 5), we observed similar particle decay trends across the decay trials between the two particle instruments. Detailed particle concentration summary statistics and timeseries from TSI 3330 and PurpleAir Classic can be found in Supplementary Information section 5.

**Figure 5.**
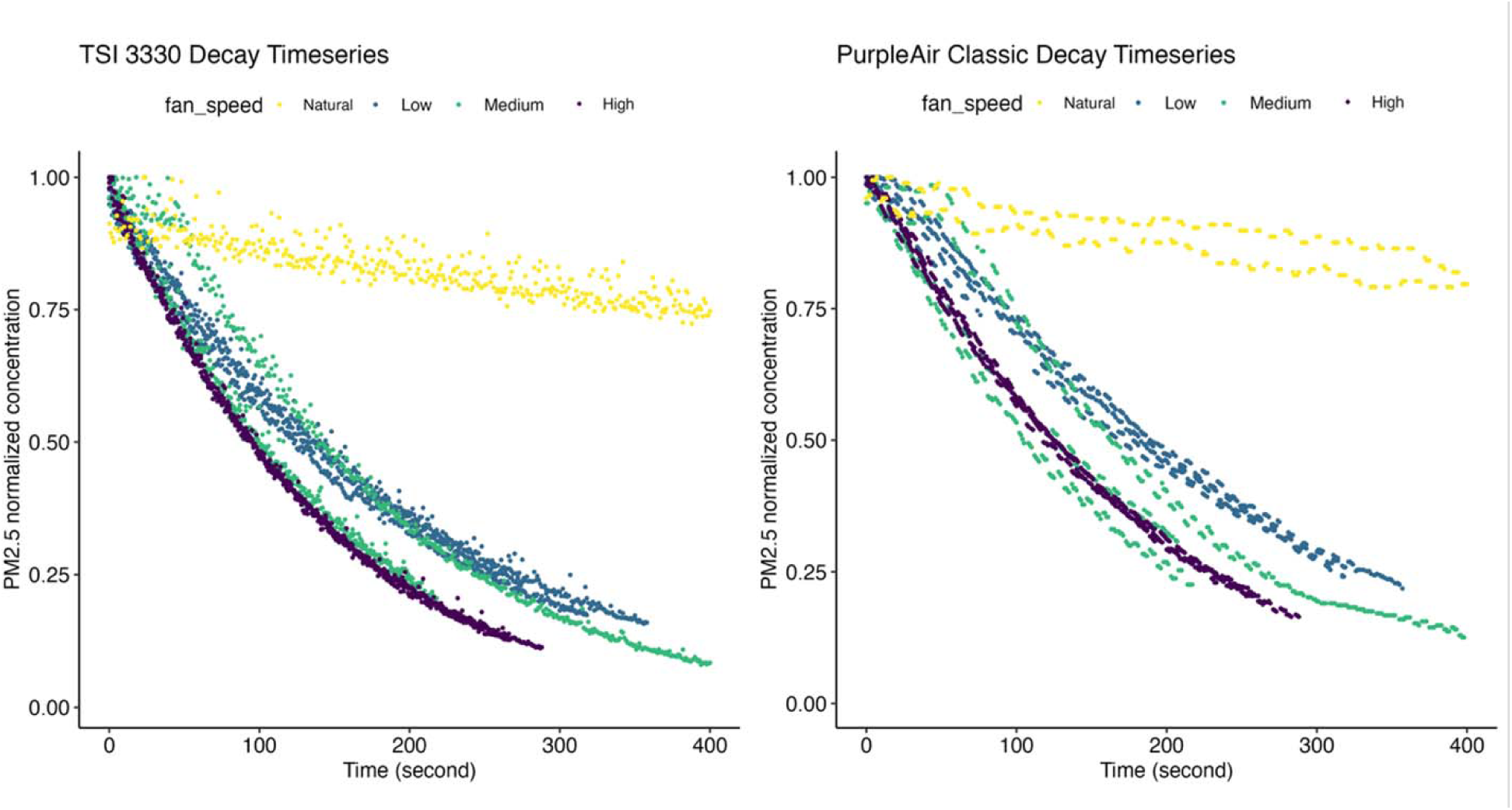
Normalized particle decay timeseries plots from TSI 3330 (left) and PurpleAir Classic (right) with each CADR box fan filter decay trial. Initial concentrations for every decay, including the natural decay, were approximately 100 μg/m^3^.

Particle decays, including natural decay, were calculated using TSI 3330 and PurpleAir Classic and the results were shown in Table 3. The natural decay constants were 1.8 (SE = 0.01) /h from TSI 3330 and 1.7 (SE = 0.01) /h from PurpleAir Classic. Decay constants for experimental trials were adjusted by the natural decay constant and were calculated from both TSI 3330 and PurpleAir Classic. CADRs for fan speeds low, medium, and high as measured by the TSI 3330 were 92.2 cfm, 129.9 cfm, and 145.2 cfm, respectively, and were 79.2 cfm, 130.1 cfm, and 116.1 cfm as measured by the PurpleAir Classic. While there were slight differences in the CADR results between the TSI 3330 and PurpleAir Classic, they generally followed the same trend of increasing CADR with higher fan speeds, particularly between low fan speed versus the other two higher fan speeds (Table 3).

**Table 3.** Summary characteristics of DIY box fan filter and particle removal efficacy at different fan speeds. “SE” columns are standard errors that accounted for uncertainty in repeated measures in decay constants from both particle instruments. Decay constants were adjusted by natural decay constant.

| Fan speed | Decay Constant,<br>/h (SE)<br>TSI 3330 | Decay Constant,<br>/h (SE)<br>PurpleAir<br>Classic | CADR, cfm<br>TSI 3330 | CADR, cfm<br>PurpleAir<br>Classic |
| --- | --- | --- | --- | --- |
| Low | 18.8 (0.06) | 15.9 (0.07) | 92.2 | 79.2 |
| Medium | 25.2 (0.09) | 25.1 (0.22) | 129.9 | 130.1 |
| High | 27.4 (0.09) | 22.6 (0.09) | 145.2 | 116.1 |

### 3.2 Community-engaged event and live demonstration

On the day of the community-based event, we used roughly 40 minutes to set up the tent exposure chamber and the DIY box fan filter particle removal experiment live demonstration. The equipment was powered by a portable solar battery pack (Jackery Portable Power Station Explorer 300; ASIN: B082TMBYR6) provided by a community partner. Around 15 community attendees stopped by for about 30 minutes and showed great interest in engaging in the live demonstration and conversations on air quality, including how to reduce exposure during WFS events, and how to build and use the DIY box fan filter. Some attendees offered to introduce the woodsmoke into the tent exposure chamber, while others closely monitored particle concentration change with the DIY box fan filter on and off through the modified Dylos DC1700 particle counter-laptop interface, where the peaks reflected the smoke introduction and the decrease in concentration was from turning on the DIY box fan filter (Fig. 6). The event organizer and attendees expressed their appreciation of being a part of the live demonstration and found the visualization of the particle concentration reduction from having DIY box fan filter in the chamber very informative. Many attendees took home a box fan and filter at the event and shared that they would use the DIY box fan filter because they now know how useful they can be during future wildfire smoke events.

**Figure 6.**
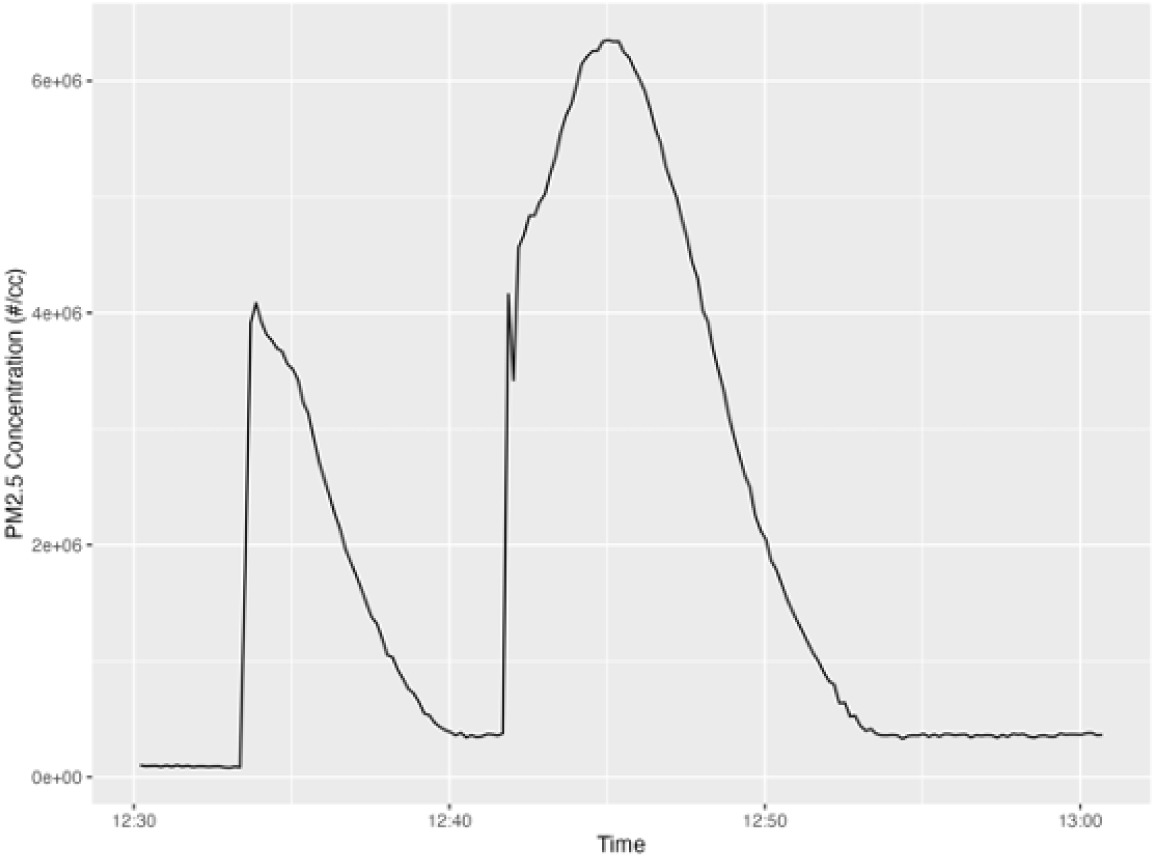
Real-time PM_2.5_ particle number concentration (Dylos) timeseries from two trial of the live demonstration.

## 4. Discussion

The tent exposure chamber provided great versatility compared to a standard AHAM exposure chamber (42,77), and allowed for experiments and demonstrations in both laboratory-based and community-based scenarios, respectively. Its portability and modular setup enabled us to transfer laboratory-based experiments to a community setting where the findings could be immediately communicated in a visual and experiential manner to complement the distribution of box fan filter supplies. The relative low-cost and commercial-off-the-shelf availability of the tent and low-cost particle sensors also allow us to easily repeat these demonstrations in diverse community settings impacted by wildfire smoke or other indoor particulate air quality challenges.

Positive feedback from the community event attendees was very encouraging. The amount of curiosity, willingness to interact and engage actively in experiments, and excitement after learning indicate that the live demonstration was successful. The tent exposure chamber realized our goals of translating lab-based experiments to community settings as an engagement tool with community members. It was particularly important to us to reach a rural agricultural population in Central Washington that experiences more frequent wildfire smoke events, but may not have previously considered air filtration as an effective strategy to improve indoor air quality during major smoke episodes, and who may not have the opportunity to view such experiments or demonstrations at the university laboratory setting in Seattle (more than 2 hours away driving) (34,35).

Further, our results using TSI 3330 and PurpleAir Classic particle instruments revealed similar DIY box fan filter CADR ratings when compared to results from previous EPA study findings (42). Holder et al. measured CADR for a variety of DIY air cleaners, including a unit made of a box fan with a MERV 13 filter (“baseline” model), in a method comparable to AHAM certification testing condition. Their results (“baseline” CADRs (cfm) at low, medium, and high fan speeds: 79.7, 96.9, and 111.2) were similar to those we measured with the low-cost exposure chamber and the low-cost PurpleAir Classic sensor (79.2 cfm to 130.1 cfm for different fan speeds). Discrepancies could be due to the limitations of the low-cost sensors, as well as due to experimental errors (e.g. tent exposure chamber not zipped well, smoke not well mixed, differences in smoke composition) for some experimental trials, or differences between our box fans and MERV 13-rated filters. Despite the variations in CADR results, the similarity between our findings for DIY box fan filter testing suggests that the tent exposure chamber can serve as a lower-cost and less complex alternative exposure chamber to other exposure chamber designs (77) (42). Moreover, although there were slight differences in the CADR results determined by the TSI 3330 research-grade particle instrument and PurpleAir Classic (which may be due to particle sizing and density assumption differences between instruments), the low-cost PurpleAir Classic was still capable of assessing clear particle decays in woodsmoke concentration for CADR quantification.

One simple engineering modification to the particle instruments likely contributed to greater interest and appreciation for the live demonstration at the community event was the interfacing of the Dylos DC1700 to a laptop. While many direct-reading particle instruments can display current numeric concentrations in real-time, most, including the TSI 3330, PurpleAir Classic, and the Dylos DC1700, do not have a display that can show the time-series of changing particle concentrations. Because of the lack of a time-series display, the decay in particle concentration from operating a PAC cannot be seen. For this reason, we used the digital serial interface on the Dylos DC1700 to send data to a laptop running a programming script to plot the concentration time-series. Being able to visualize real-time particle concentration reduction due to DIY box fan filter likely promoted attendees willingness to take a DIY box fan filter home to use for future wildfire smoke events.

A few limitations were observed during the CADR testing experiment and the community-based live demonstration. First, the use of low-cost particle sensors may not provide accurate absolute results, but because the decays and CADR are based on relative concentration changes over time, accurate absolute concentrations are less important, which was confirmed by observing similar CADR findings compared to experiments conducted under AHAM’s certified testing condition (42). Moreover, we used a relatively simple approach to generate wood smoke as a particle source that can be difficult to produce consistently for experiments. But engineered particle sources (e.g., Arizona test dust, ASHRAE dust) could also be used for similar future experiments. Low-cost particle sensors, due to their particle sensing technology and data logging limitations (e.g., low logging rate), can present challenges for experimental studies. However, we were fortunate in our case, that there were manufacturer-supported recommendations for modifying the PurpleAir Classic to log measurement data at faster intervals, and the availability of two sensors within the PurpleAir Classic to confirm observed decays were consistent between both sensor channels. Nevertheless, researchers using low-cost particle sensors should be aware of the sensors’ recommended use cases/applications, and any limitations (e.g., temporal filtering) that may affect experimental protocols that require measurement of concentration decays over short time periods. Lastly, despite the success of the community-based event in Central Washington, future researchers planning to bring the tent exposure chamber system to perform a live demonstration should be aware of weather conditions forecast for the time of the event. Even with the mylar-lined canvas and sturdy metal frame, the tent may not be an ideal exposure chamber under extreme weather conditions (e.g., rain, snow, strong winds).

## 5. Conclusion

Overall, the tent exposure chamber was found to be useful in both laboratory-based and community-based settings, specifically in the evaluation and demonstration of DIY box fan filters. Through the community-based event, feedback from attendees confirmed the importance and usefulness of the novel tent exposure chamber system and transferring the experiments from the university laboratory to a community demonstration. Future researchers may consider similar equipment and strategies to bring science to communities and encourage visualization and interaction between the research members and community partners to promote awareness, knowledge, and behavioral change that protect members of communities from wildfire smoke.

## List of Abbreviations

- WFS: wildfire smoke
- PHSKC: Public Health Seattle & King County
- WALNI: Washington State Labor and Industries
- CHF: congestive heart failure
- COPD: chronic obstructive pulmonary disease
- PAC: portable air cleaner
- PM_2.5_: fine particulate matter with an aerodynamic diameter smaller than 2.5 μm
- HEPA: high efficiency particulate air
- DIY: do-it-yourself
- EPA: Environment Protection Agency
- CADR: clean air delivery rate
- AHAM: Association of Home Appliance Manufacture
- UW: University of Washington
- CEHE: Center for Environmental Health Equity
- TCTAC: Thriving Communities Technical Assistance Center
- DEOHS: Department of Environmental and Occupational Health Sciences
- CDC: Centers for Disease Control and Prevention
- CSV: comma-separated-value
- AQI: air quality index
- SD: standard deviation
- SE: standard error
- CTEK: Cultural Traditional Ecological Knowledge Planner

## Declaration

- Ethics approval and consent to participate: University of Washington Human Subject Division reviewed the Institutional Review Board Application (STUDY00025081) on March 5, 2026, and determined the described activities in this manuscript were not research.

- Consent for publication: not applicable

- Availability of data and materials: data and materials are available in supplementary information; raw data and R scripts are available upon request

- Competing interests: the author declared no competing interests

- Funding: Research reported in this publication was supported by the University of Washington EDGE Center of the National Institutes of Health under award number: P30ES007033. Funders had no role in the design of the study, collection, analysis, interpretation of data, or in manuscript preparation.

- Authors’ contributions: Lilian Liu: Conceptualization, Methodology, Formal analysis, Investigation, Data curation, Writing – Original draft. Shirley Huang: Experiment design, Validation, Writing - Review & Editing. Alison Hirata: Experiment design, Writing - Review & Editing. Ningrui Liu: Writing - Review & Editing. Jeffry Shirai: Writing - Review & Editing. Chris Zuidema: Writing - Review & Editing. Elena Austin: Validation, Writing - Review & Editing. Edmund Seto: Conceptualization, Methodology, Data curation, Writing - Review & Editing.

## Data Availability

All data produced in the present study are available upon reasonable request to the authors

## Acknowledgements

We thank our community partner in Central Washington for their enthusiasm and invitation to the event. KHIMSTONIK would like to express a special thank you to the Yakama Nation Tribes and community members that were able to participate with our initiative.

