## Supplementary Information for "Translating a novel wildfire smoke exposure chamber system from lab-based experiments to community-engaged activities"

1. **Tent exposure chamber detail**

The tent is 120 inches (3.05 m) in length, 60 inches (1.52 m) in width, and 80 inches (2.03 m) in height. Two major components that set up the tent are the metal frame and mylar-lined Oxford canvas. A product information diagram can be found on the [retailer website](https://www.amazon.com/VIVOSUN-Strengthened-Reflective-Hydroponics-VSFD6500%C3%972/dp/B0CGHJMJGT/ref=sr_1_6?dib=eyJ2IjoiMSJ9.hmITYNpgnGUdWPrgXLp_Ti4_9thZkO4_sLQ-qK0fUH_YRLt36Eo_SwYV5tQReWpKRqLJuIORqtBLxtR0V-0VDSnHTbz-H4b-g3JRpEWyYOffhtJf0c_rp-Qk4-OUVQgl9VeTRC2ZSaMsnS1E8Cc-gg6vze21ABaSIkj1UYA6pgNAWeMDg01Zpig5X_dpImjsJ25rnMu1kb2oHdw7DjN51pFsmA1D-7gNAs39yyVYBIvz0F4AkO4wORJmewXKSMCv549hm5LSAeLH4a8RdFxlgovhXZNX18dZKuwQCp4jcJQ.w96IJCK-bNhfWsgiL8qyRXuhXmvxY_wiYVDRmzypZzE&dib_tag=se&keywords=vivosun%2Bgrow%2Btent%2B80%2B60%2B120&qid=1769207975&sr=8-6&th=1) where the instruction sheet shows the details of the tent construction. Briefly, the metal frame contains a floor (6 A poles, connected by 2 three-way T-shaped connectors and 4 three-way elbow corner connectors), a roof (7 A poles, connected by 2 four-way elbow connectors and 4 three-way elbow corner connectors), and 4 walls (6 units of vertical poles made of B and C poles). The tent has 4 doors (2 on the front wall and 2 at the back wall) that can open/close via zip linings and 2 plastic windows on the front wall. The tent came with an extra floor lining to provide extra protection, which users can use upon their discretion.

2. **Woodsmoke generation protocol**

To increase the particle concentration inside the tent exposure chamber, we burned approximately 1 tablespoon of fine woodchip in the smoking gun and introduced smoke into the tent through unzipping the tent door. A handheld lighter was used to light the woodchip, and all woodchips must be completely lit/burned before finishing the smoke introduction. TSI 3330 was used to monitor the particle concentration in real-time. For each decay trial, target particle concentration was 100 μg/m^3^. When the concentration was too high (e.g. 500 μg/m^3^), we opened the tent and allow the smoke particle to exit the tent exposure chamber until it returned to around the target concentration. When the concentration was too low (e.g. 50 μg/m^3^), we will burn more woodchips until the concentration reached around the target concentration.

3. **DIY box fan filter face velocity testing**

A MERV13-rated filter was attached to the inlet of a Lasko 20 in. 3 Speeds Box Fan with duct tape. A 1-foot cardboard shroud was attached to the 4 sides of the DIY box fan filter at the side of the inlet. TSI 9565 VELOCICALC Air Velocity Meter was used to test the box fan face velocity at 9 locations on the fan (Fig. S1), which was later used to calculate the air flow rate (Eq. 1). A total of 81 measurements were taken for 3 fan speeds at 9 different locations. Final face velocities at 3 fan speeds were the averages from all repeated measurements. DIY box fan filter for face velocity testing with a shroud is shown in Fig. S2.

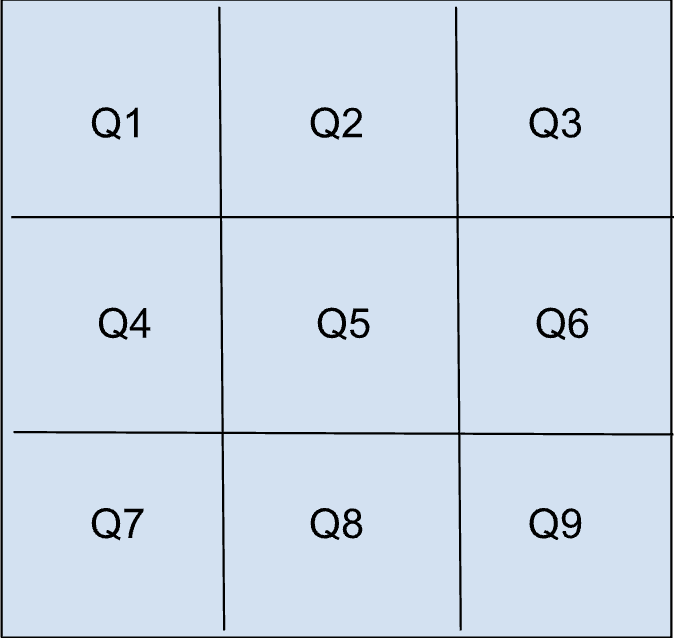

Figure S1. DIY box fan filter face velocity measurement locations. Each measurement took place about ½ foot away from the fan inlet face. Each location at one fan speed were measured 3 times. Three fan speeds (low, medium, high) were all measured for all 9 locations. A total of 81 measurements were recorded using TSI 9565.

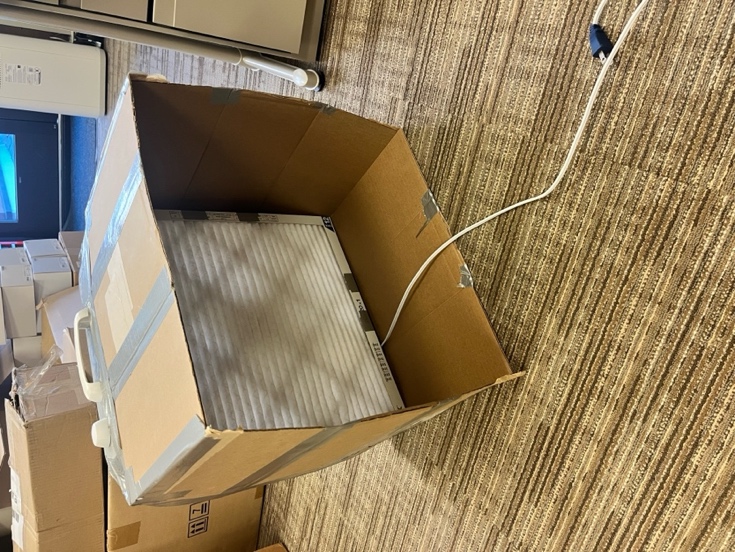

Figure S2. Photo of the DIY box fan filter with a shroud made of card box for fan face velocity and air flow testing under ambient indoor environment.

4. **Center for Environmental Health Equity Handouts**

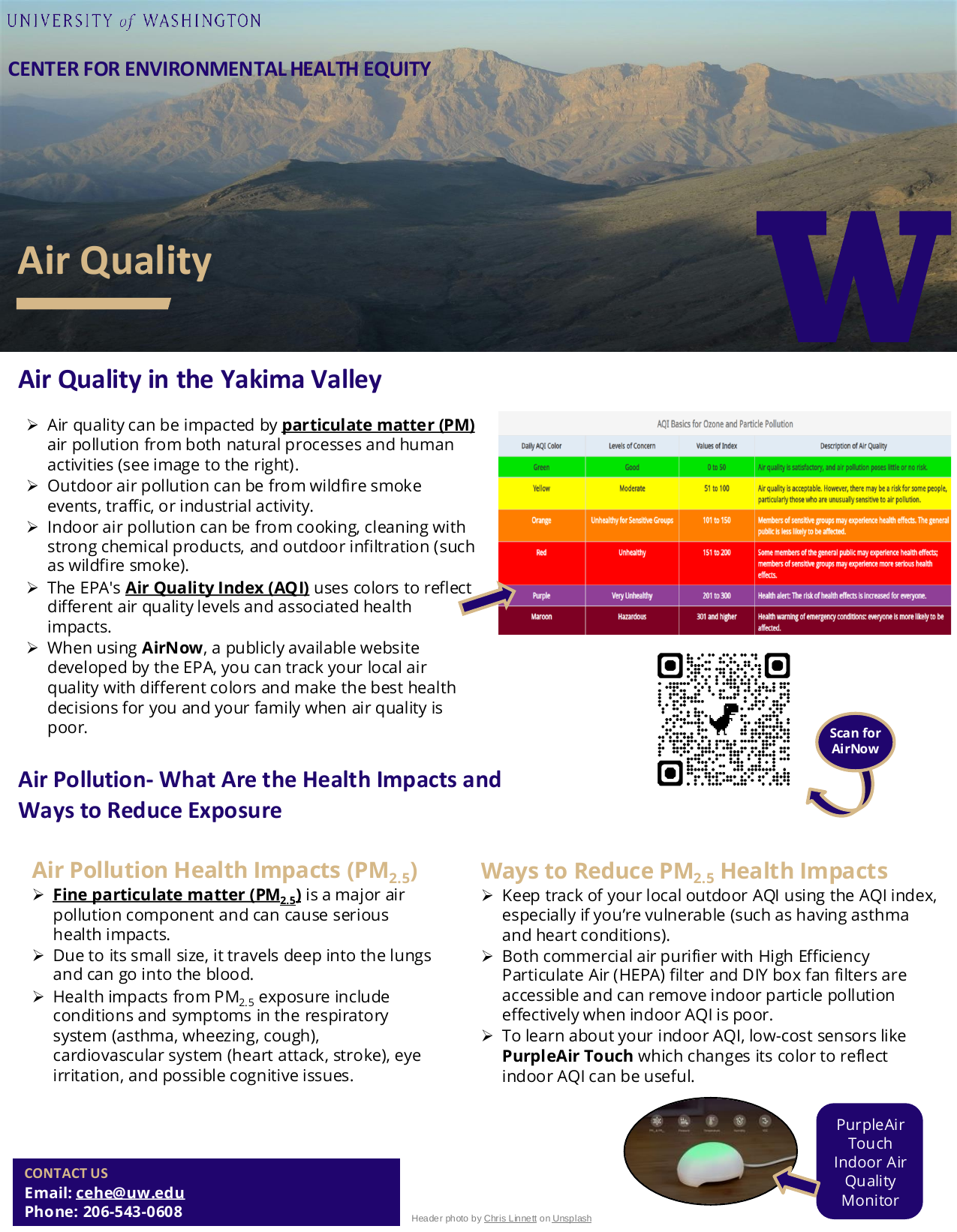

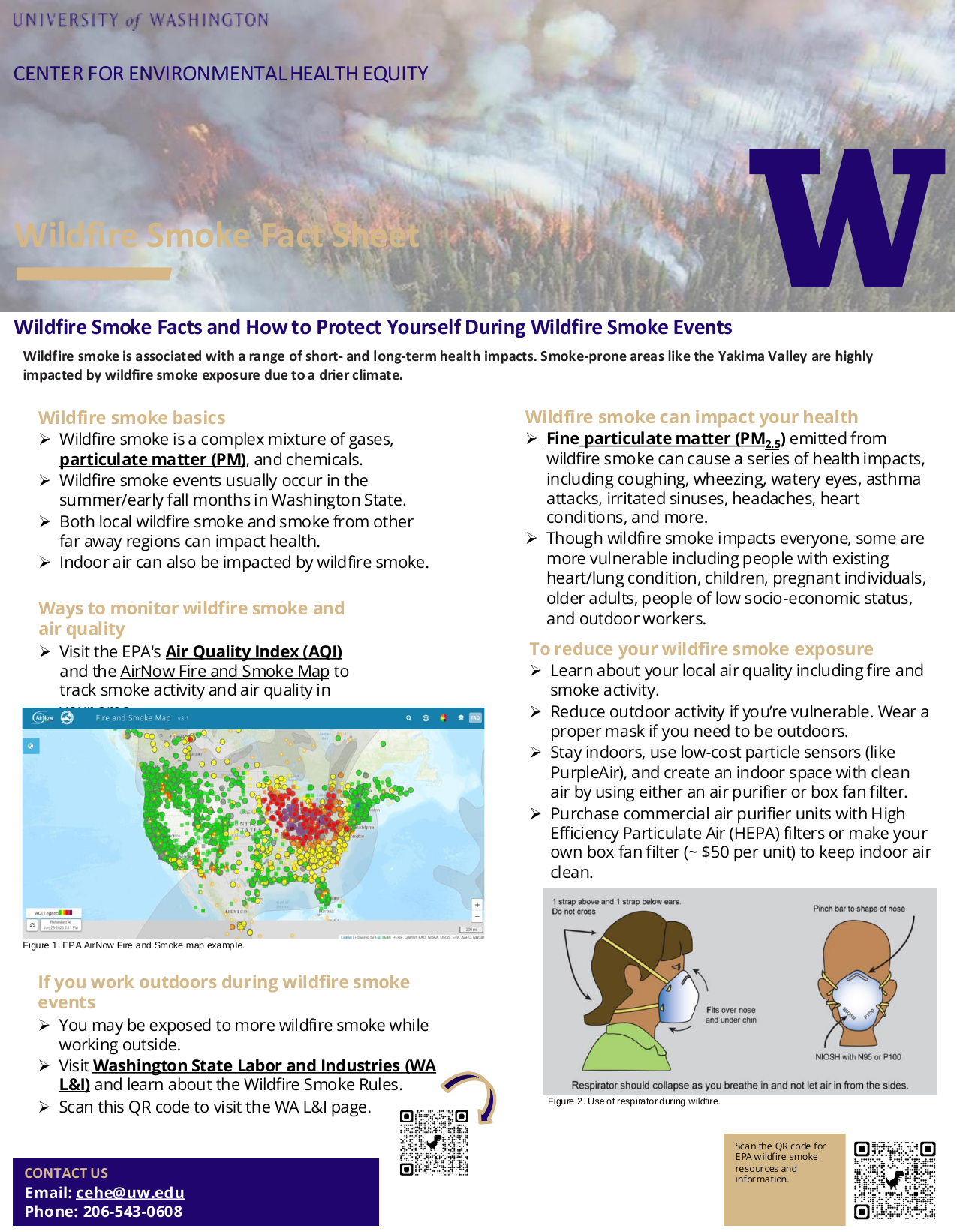

5. **DIY box fan filter CADR testing**

To increase the particle concentration inside the tent exposure chamber, we burned approximately 1 tablespoon of fine woodchip in the smoking gun and introduced smoke into the tent through unzipping the tent door. A mixing fan inside the tent exposure chamber was turned on at the highest speed during the entire experiment to homogenize inside air. Once TSI 3330 total particle concentration read around 100 μg/m^3^, we zipped the tent and turned on the DIY box fan filter. When TSI 3330 concentration dropped to around 10 μg/m^3^, we turned off the DIY box fan filter. Low, medium, and high speeds were each was tested 3 times for 9 quadrants of the fan. 27 sets decay were collected for CADR analysis (Eq. 2) with decay constants derived from the tent experiment trials (Eq. 3).

6. **Summary statistics and plots from lab-based CADR testing**

Table S1. PM_2.5_ mass concentration (μg/m^3^) summary statistics from TSI 3330 during DIY box fan filter particle removal efficacy experiment at UW. SD stands standard deviation.

| Fan speed/experiment trial | PM_2.5_ min | PM_2.5_ max | PM_2.5_ median | PM_2.5_ mean | PM_2.5_ SD |
| --- | --- | --- | --- | --- | --- |
| Low_1 | 13.04 | 82.8 | 33.8 | 38.21 | 19.44 |
| Low_2 | 10.93 | 62.81 | 25.25 | 28.95 | 13.84 |
| Low_3 | 14.73 | 64.23 | 31.36 | 33.87 | 13.87 |
| Medium_1 | 0.83 | 75.28 | 4.34 | 14.94 | 20.29 |
| Medium_2 | 12.42 | 60.42 | 27.79 | 31.1 | 13.98 |
| Medium_3 | 13.81 | 60.29 | 29.66 | 32.05 | 13.28 |
| High_1 | 9.65 | 76.06 | 27.83 | 32.85 | 18.81 |
| High_2 | 10.25 | 91.86 | 32.1 | 38.03 | 22.86 |
| High_3 | 12.34 | 88.31 | 34.39 | 39.14 | 21.06 |
| Natural Tent Decay | 28.2 | 56.88 | 38.6 | 39.29 | 6.73 |

Table S2. PM_2.5_ mass concentration (μg/m^3^) summary statistics from PurpleAir Classic during DIY box fan filter particle removal efficacy experiment at UW. SD stands standard deviation.

| Fan speed/experiment trial | PM_2.5__min | PM_2.5__max | PM_2.5__median | PM_2.5__mean | PM_2.5_ SD |
| --- | --- | --- | --- | --- | --- |
| PA_Low_1 | 86 | 394 | 204 | 219 | 95 |
| PA_Low_2 | 113 | 469 | 230 | 256 | 104 |
| PA_Low_3 | 144 | 432 | 268 | 277 | 88 |
| PA_Medium_1 | 1 | 263 | 31 | 64 | 78 |
| PA_Medium_2 | 58 | 257 | 126 | 138 | 59 |
| PA_Medium_3 | 138 | 449 | 231 | 272 | 99 |
| PA_High_1 | 96 | 504 | 238 | 259 | 120 |
| PA_High_2 | 84 | 510 | 223 | 251 | 129 |
| PA_High_3 | 104 | 510 | 248 | 269 | 119 |
| PA_Natural Tent Decay | 97 | 177 | 131 | 132 | 22 |

Table S3. PM_2.5_ mass concentration (μg/m^3^) summary statistics from Dylos DC1700 during DIY box fan filter particle removal efficacy experiment at UW.

| Fan speed/  experiment trial | PM_2.5__min | PM_2.5__max | PM_2.5__median | PM_2.5__mean |
| --- | --- | --- | --- | --- |
| Dylos_Low_1 | 2837 | 5884 | 3437 | 749 |
| Dylos_Low_2 | 1890 | 2872 | 2603.5 | 243 |
| Dylos_Low_3 | 2473 | 3236 | 2885 | 179 |
| Dylos_Medium_1 | 289 | 8045 | 982.5 | 2358 |
| Dylos_Medium_2 | 2912 | 4632 | 3882.5 | 365 |
| Dylos_Medium_3 | 1990 | 2656 | 2442 | 178 |
| Dylos_High_1 | 1509 | 4078 | 2652 | 539 |
| Dylos_High_2 | 1906 | 7963 | 2849 | 1309 |
| Dylos_High_3 | 2345 | 7501 | 3283 | 1139 |
| Dylos_Natural Tent Decay | 5676 | 6495 | 6020 | 154 |

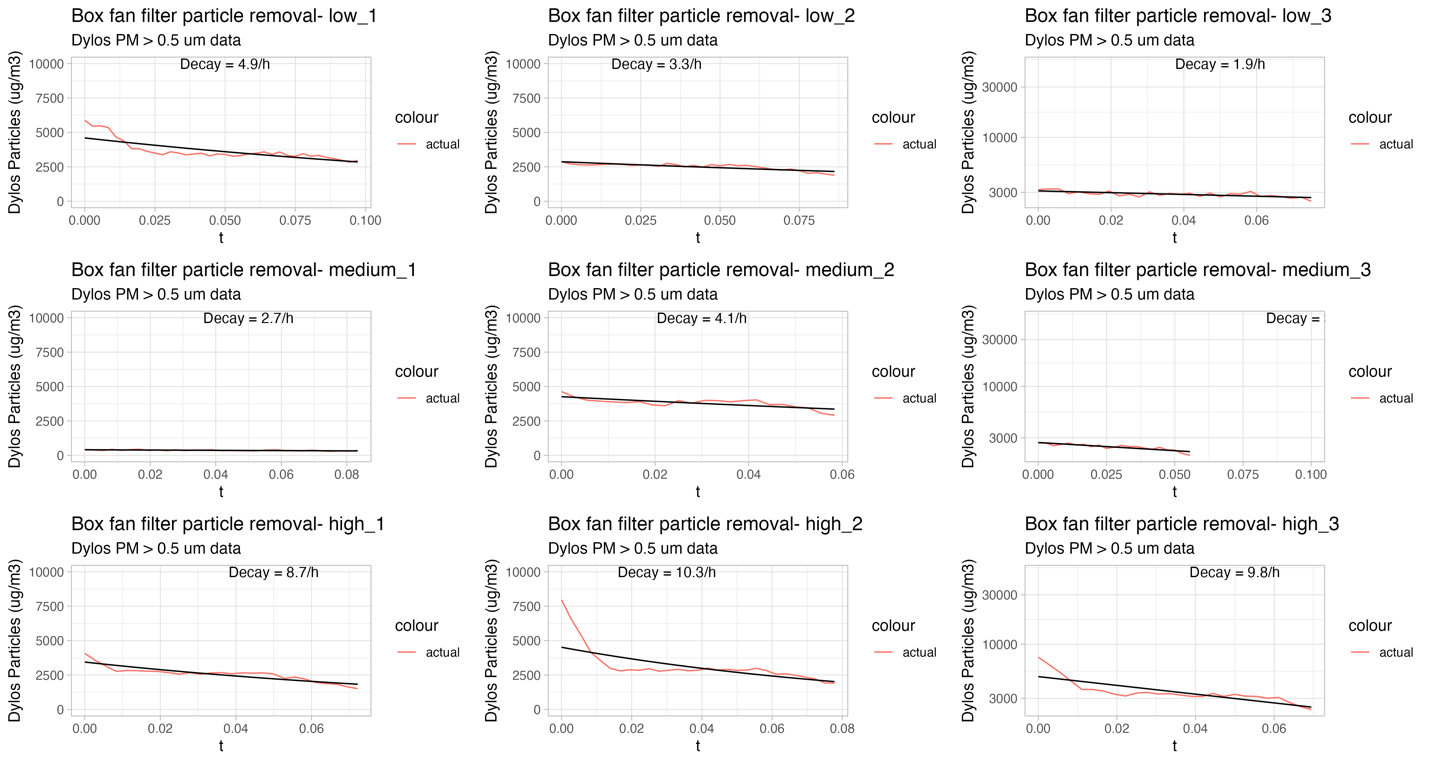

Figure S3. Dylos DC1700 PM_2.5_ mass concentration (μg/m^3^) decay timeseries with decay constant.

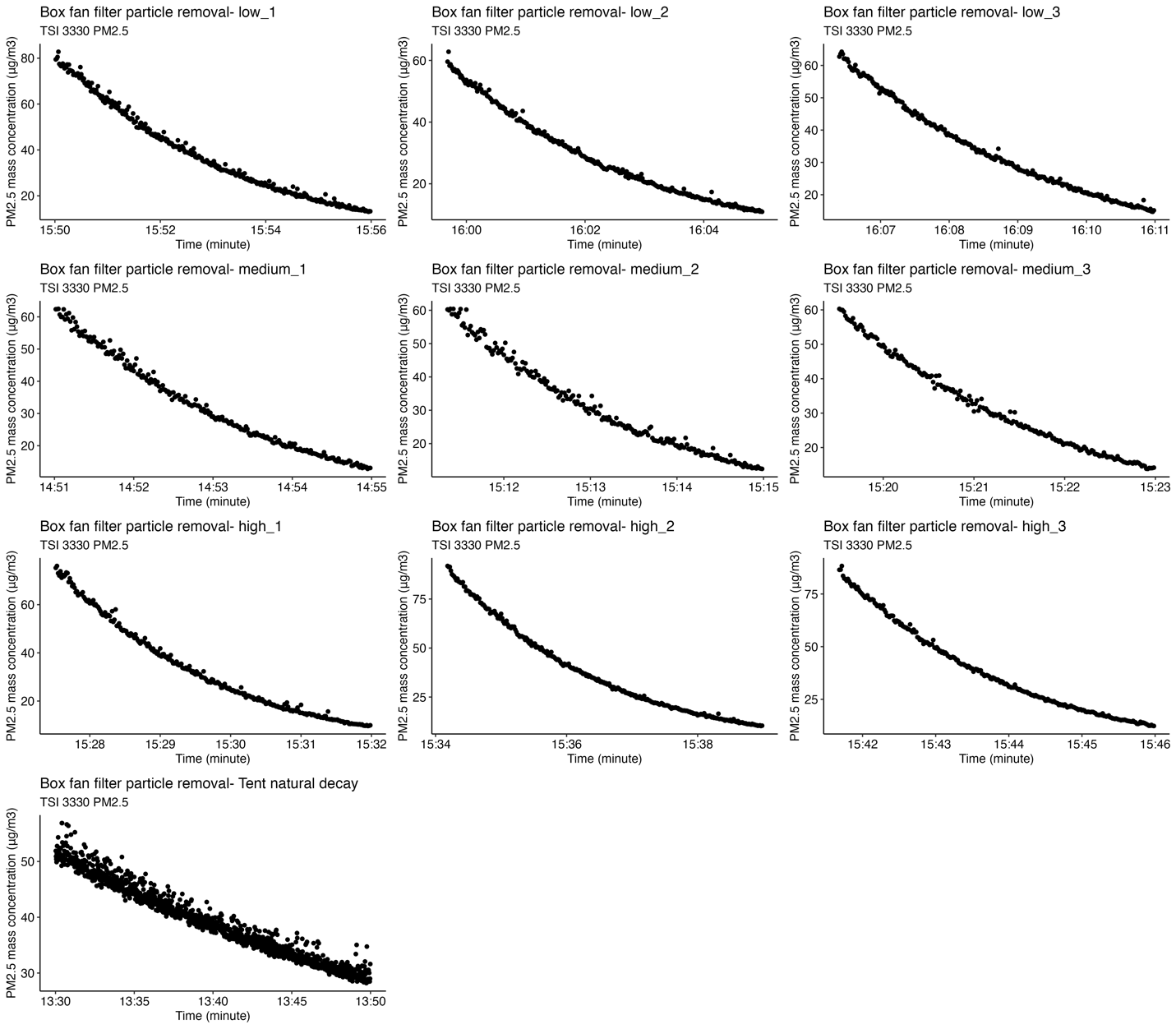

Figure S4. Time-series plots from TSI 3330 for DIY box fan filter decay trials at different experimental stages.

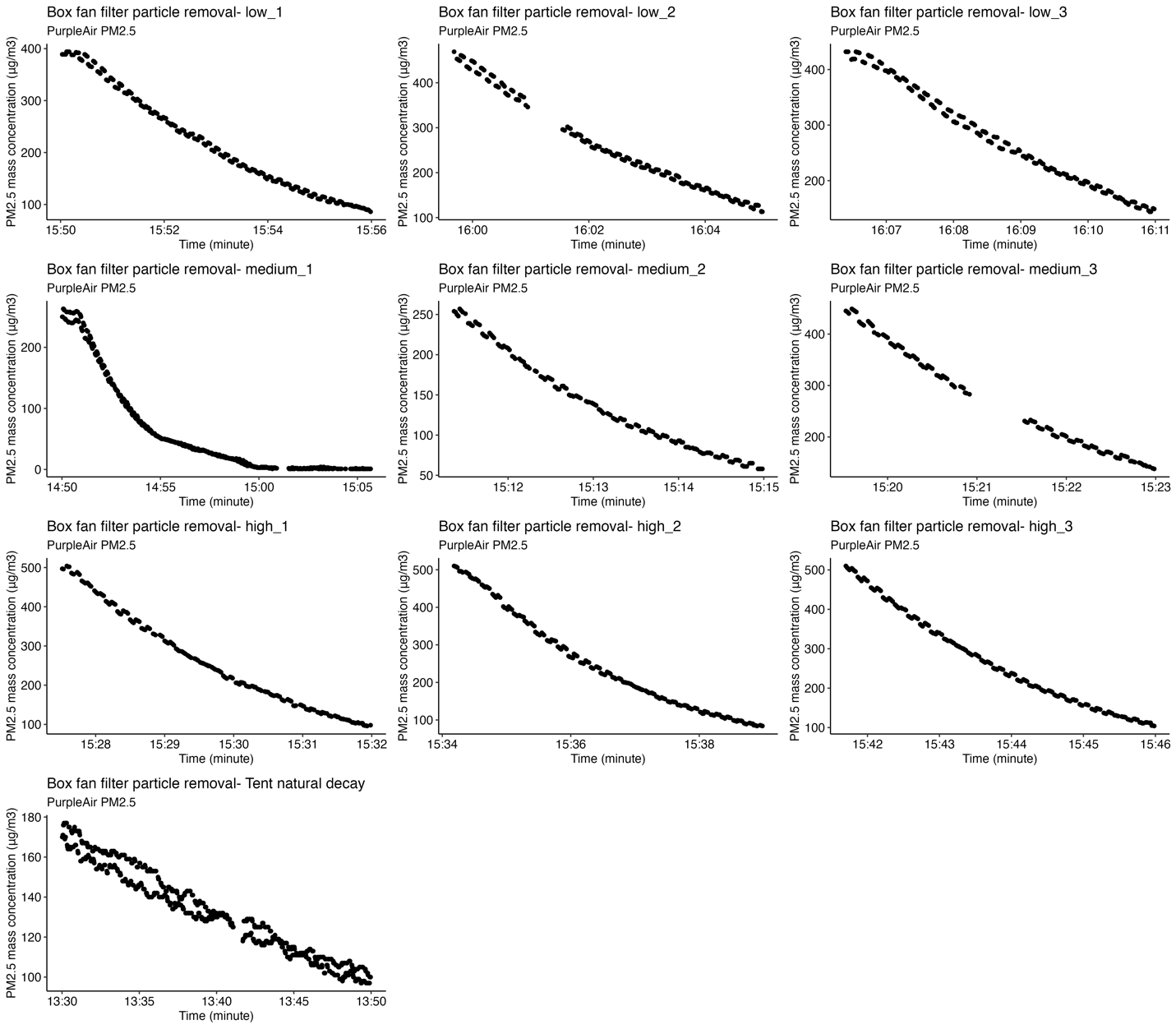

Figure S5. Time-series plots from the two particle sensors in the PurpleAir Classic for DIY box fan filter decay trials at different experimental stages. Note, PurpleAir Classic has 2 particle sensors that work simultaneously. The timeseries plots would appear to have two parallel lines when the particle concentration results from two sensors differed slightly.

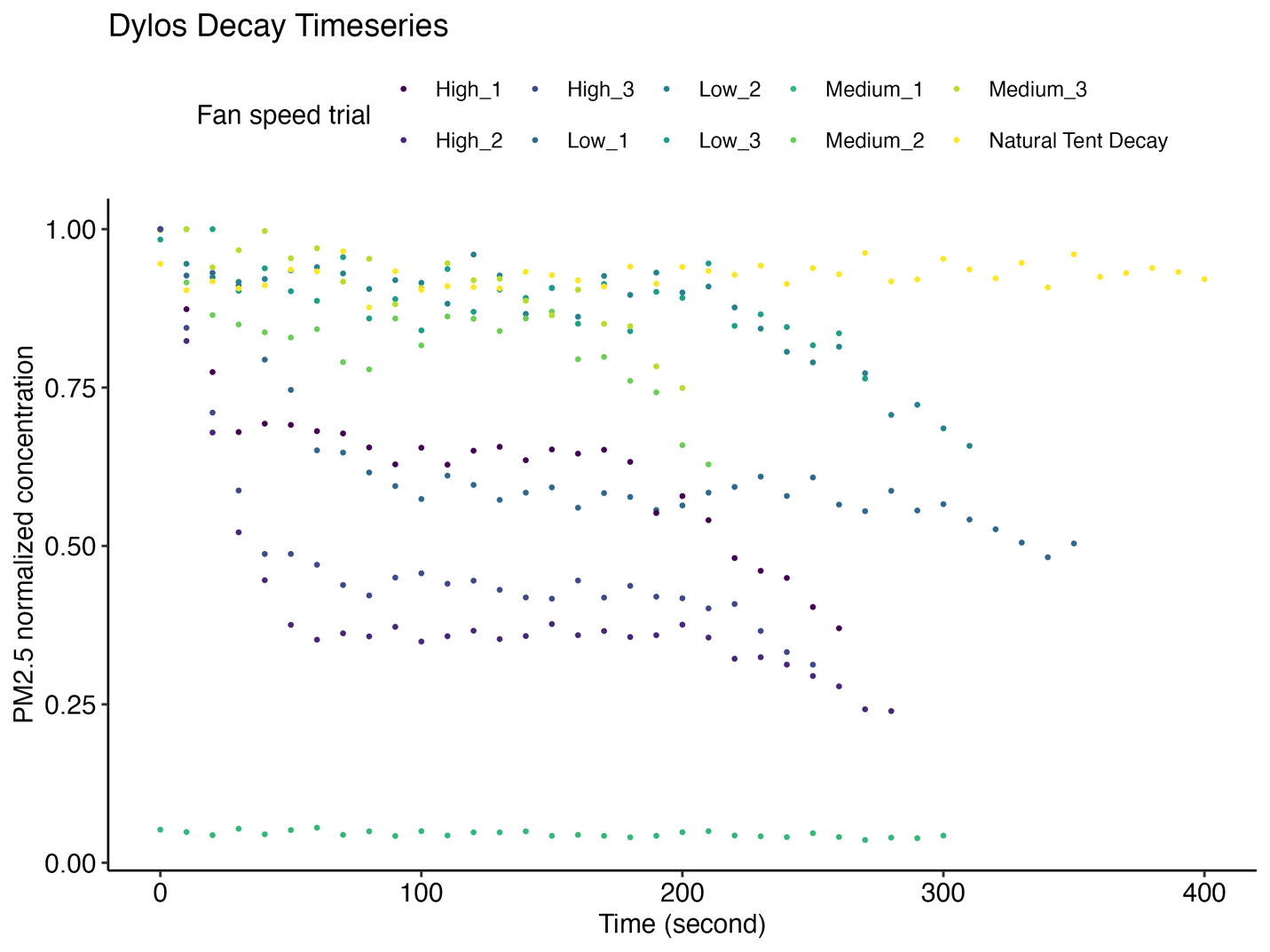

Figure S6. Normalized particle decay timeseries plots from Dylos DC1700 with each CADR box fan filter decay trial. Initial concentrations for every decay, including the natural decay, were approximately 100 μg/m^3^.
